# Independent and joint effects of genomic and exposomic loads for schizophrenia on distressing and persisting psychotic experiences in adolescence

**DOI:** 10.1101/2024.11.11.24316985

**Authors:** Matteo Di Vincenzo, Thanavadee Prachason, Gaia Sampogna, Angelo Arias-Magnasco, Bochao Danae Lin, Lotta-Katrin Pries, Jim van Os, Bart P.F. Rutten, Ran Barzilay, Andrea Fiorillo, Sinan Guloksuz

**Affiliations:** Department of Psychiatry, University of Campania “Luigi Vanvitelli”, Naples, Italy; Department of Psychiatry, Faculty of Medicine Ramathibodi Hospital, Mahidol University, Bangkok, Thailand; Department of Psychiatry and Neuropsychology, School for Mental Health and Neuroscience, Maastricht University Medical Centre, Maastricht, The Netherlands; Department of Psychiatry, UMC Utrecht Brain Centre, University Medical Centre Utrecht, Utrecht University, Utrecht, The Netherlands; Department of Psychosis Studies, Institute of Psychiatry, Psychology & Neuroscience, King’s College London, London, UK; Department of Psychiatry, Perelman School of Medicine, University of Pennsylvania, Philadelphia, Pennsylvania; Lifespan Brain Institute of the Children’s Hospital of Philadelphia and Penn Medicine, Philadelphia, Pennsylvania; Department of Child and Adolescent Psychiatry and Behavioral Science, Children’s Hospital of Philadelphia, Philadelphia, Pennsylvania; Department of Psychiatry, Yale University School of Medicine, New Haven, Connecticut

**Keywords:** psychotic experiences, polygenic risk score, environment, schizophrenia, gene-environment interaction, adolescence

## Abstract

To assess the longitudinal associations of genomic and exposomic liabilities for schizophrenia, both independently and jointly, with distressing psychotic experiences (PEs) and their persistence in early adolescence. The Adolescent Brain and Cognitive Development Study data from children with European ancestry were used (n=5,122). The primary outcome was past-month distressing PEs at 3-year follow-up. Secondary outcomes were lifetime distressing PEs defined at varying cutoffs of persistence (from ≥ 1-4 waves). Multilevel logistic regression models were used to test the independent and joint associations of the binary modes (risk-category defined as above 75th percentile) of polygenic risk score for schizophrenia (PRS-SCZ_75_) and exposome score for schizophrenia (ES-SCZ_75_) on the outcomes. The relative excess risk due to interaction (RERI) was determined using the delta method to indicate departure additive interaction. PRS-SCZ_75_ was statistically significantly associated with lifetime distressing PEs (≥ 1 wave) (OR 1.29 [95% CI 1.08, 1.53]) and repeating distressing PEs ≥ 2 waves (OR 1.34 [95% CI 1.08, 1.65]) but not with past-month distressing PEs or repeating distressing PEs at a higher cutoff of persistence. ES-SCZ_75_ was consistently associated with past-month and repeating distressing PEs at all cutoffs, with increasing strength of association as a function of PEs persistence (one wave: OR 2.77 [95% CI 2.31, 3.31]; two waves: OR 3.16 [95% CI 2.54, 3.93]; three waves: OR 3.93 [95% CI 2.86, 5.40]; four waves: OR 3.65 [95% CI 2.34, 5.70]). There was evidence for additive interaction between ES-SCZ_75_ and PRS-SCZ_75_ for lifetime distressing PEs (RERI=1.26 95%CI: 0.14, 2.38), and repeating distressing PEs ≥ 2 waves (RERI=1.79, 95%CI: 0.35, 3.23). Genomic and exposomic liabilities for schizophrenia were independently and jointly associated with distressing PEs, as well as their persistence in early adolescence.

## 1. Introduction

Psychotic experiences (PEs) are observed in around 10% of children and adolescents and increase the odds of having any mental and psychotic disorders by three- and four-fold, respectively^1,2^. Specifically, PEs causing distress (i.e., distressing PEs) are associated with developmental delay, neuroimaging findings such as reduced subcortical and cortical volumes, and suicidal behavior^3,4^. Despite the transient nature of most PEs, a recent meta-analysis showed that around one-third of people having PEs at baseline reported a second PE within a year^5^. The persistence of PEs, especially when related to distress, is also linked with multiple mental disorders and greater impairments^4,5^. Given their clinical significance as a marker for later clinical syndromes, a better understanding of the etiology of distressing PEs as well as their persistence among youth is needed.

The shared heritability of PEs with psychotic disorder has long been discussed^6^. Genome-wide association studies (GWAS) have provided further evidence for shared genetic liability between PEs and schizophrenia^7,8^. In recent years, polygenic risk score (PRS), generated by summing the risk alleles weighted by their effect sizes derived from related GWAS, has been increasingly utilized to estimate an individual’s genetic predisposition to behavioral phenotypes and psychiatric disorders. Prior studies have shown that PRS for schizophrenia (PRS-SCZ) is associated with subclinical psychosis expression in children and adolescents^9–11^ as well as adults^12^. However, since other researches did not obtain similar results, probably due to differences in PRS-SCZ calculation and population sample, further investigations are required^13,14^.

Similar to genetic liability, an additive risk of environmental exposures has been shown to increase the odds of psychosis expression^15,16^. From the conceptualization of “exposome”, a dense network of interdependent environmental exposures^17^, the exposome score for schizophrenia (ES-SCZ) has been developed as a predictive modeling approach^18^. It is calculated by summing the weighted risks of nine binary variables underlying well-established non-genetic risk factors for schizophrenia. Apart from its predictive ability for clinical psychosis^18^, ES-SCZ is also associated with schizotypal traits in siblings of patients with schizophrenia and healthy subjects, suggesting the interplay between the genetic and environmental risks for schizophrenia across psychosis continuum^12^.

To the best of our knowledge, no study has tested the influence of exposomic liability for schizophrenia on subclinical psychosis expression, both independently and jointly with the genomic liability, using a prospective design. Moreover, most previous studies rarely examined the persistence of distressing PEs despite its association with poor prognostic features^4,5^. Given these gaps, we aimed to assess the longitudinal associations of PRS-SCZ, ES-SCZ, and their interaction with distressing PEs as well as their influences on symptom persistence, in a general population-based sample of young adolescents.

## 2. Methods

### 2.1 Participants

A sample of 11,876 participants was derived from the Adolescent Brain and Cognitive Development (ABCD) cohort, an ongoing multisite longitudinal study conducted in the United States to assess biological and behavioral development from early adolescence to young adulthood^19^. The ABCD Data Release 5.1 (see Acknowledgements) was used, and participants of European descent with good-quality genotyping data were included in the current analyses.

The ABCD Study was approved by the Centralized Institutional Review Board (IRB) of the University of California-San Diego and by local research site IRBs^19^. Written informed consent and assent were provided by participating parents/caregivers and adolescents, respectively^19^.

### 2.2 Variables

#### 2.2.1 Psychotic experiences

PEs were assessed using the Prodromal Questionnaire-Brief Child Version (PQ-BC), a self-reported tool validated for the school-age population^20^. It comprises 21 items asking about PEs (e.g., unusual thought content, perceptual abnormality) in the past month. When adolescents reported the presence of any PEs, they were asked to rate the distress level related to the experience from 1 to 5. Consistent with previous research^11^, “distressing PEs” were regarded as present if at least one PE was rated 3 points or more; otherwise, absent. Past-month distressing PEs at 2-year follow-up were used as the primary outcome. To explore the effects of risk exposure on the persistence of distressing PEs, four dichotomous variables were generated at varying degrees of persistence of distressing PEs from baseline to 3-year follow-up as secondary outcomes: “lifetime distressing PEs” (i.e., presence of distressing PEs ≥ 1 wave), “repeating distressing Pes ≥ 2 waves” (i.e., presence of distressing PEs ≥ 2 waves), “repeating distressing PEs ≥ 3 waves” (i.e., presence of distressing PEs ≥ 3 waves), and “persisting distressing PEs” (i.e., distressing PEs in all 4 waves).

#### 2.2.2 Polygenic risk score for schizophrenia

PRS-SCZ was constructed for each participant who passed the genetic and sample quality control. The pre-imputational quality control (Table S1), relatedness and ancestry distributions (Table S2 & Table S3), post-imputational data (Table S4) and principal components analysis for ancestry (Figure S2 & Figure S5) are reported in detail in the online supplement. PRS-SCZ were built using data from the most recent schizophrenia GWASs (European subsample) based on 53,386 cases and 77,258 controls^21^.

PRS_cs-auto_-SCZ^22^ was used to infer PRS generated using posterior SNP effect sizes, by placing a continuous shrinkage (cs) prior on SNP weights reported in the latest GWAS for schizophrenia^21^ and combined with an external linkage disequilibrium reference panel, such as the 1000 Genomes Project European Sample (https://github.com/getian107/PRScs). To compute posterior effect sizes, the default settings of PRS-cs-auto were used (See online supplement). After calculation of posterior effect sizes, PRSs were calculated using ‘--score’ function and the SUM modifier in PLINK1.9. After quality control, 742,011 variants were used in the PRS calculation. Besides PRS_cs-auto_-SCZ, we also calculated PRS_ice_-SCZ by the clumping and threshold methods using PRSice2^23^ for sensitivity analysis to verify the results.

#### 2.2.3 Exposome score for Schizophrenia (ES-SCZ)

Following prior studies^12,18^, ES-SCZ was generated using nine environmental exposures: emotional neglect, physical neglect, emotional abuse, physical abuse, sexual abuse, cannabis use, winter birth, hearing impairment, and bullying. Variables in the ABCD dataset were carefully selected to generate each exposome factor aligning with the original ES-SCZ study^18^ and prior studies involving these risk factors in the ABCD dataset^24,25^. To ensure the antecedence of the exposure to the outcome measurement at 3-year follow-up, ES-SCZ was calculated from lifetime exposure to each environmental risk (0 = “absent”; 1 = “present”) using the information at baseline, 1-year follow-up, and 2-year follow-up. The variables used to construct each exposome factor are provided in Table S7, and the detailed construction of ES-SCZ is described in the online supplement. Briefly, emotional neglect was derived from the Child Report of Parent Behaviors Inventory^26^, asking children to rate their perceived levels of parents’ warmth and closeness. Physical neglect was regarded as present if children had experienced any food shortage, history of maternal or paternal alcohol use, and poor parental supervision. Emotional abuse, physical abuse, and sexual abuse were derived from the PTSD module of parent-reported Kiddie Schedule for Affective Disorders and Schizophrenia for DSM-5^27^. Cannabis use was derived from multiple assessments based on children’s reports from baseline to 2-year follow-up, including mid-year telephone interviews. Winter birth was estimated by leveraging the precise date of the interview at baseline and the child’s age in months at the time of the interview. Hearing impairment was derived from the Parent Medical History Questionnaire, asking if the child had ever been to a doctor for hearing problems. Bullying was derived from the Parent Diagnostic Interview for DSM-5 Background Items, asking if the children had any problems with bullying at school or in the neighborhood. Finally, the weighted risks (i.e., the exposure multiplied by its log odds for schizophrenia) of the nine exposures were summed up and added by 2 for ease of interpretation (see the online supplement).

#### 2.2.4 Covariates

Age in years at the time of PE assessment, sex, baseline family income, and highest parental education were included in the models as covariates. Model 1 analysis comprised only age and sex, while all covariates were included in Model 2. The detailed description of the covariates is presented in Table S7 in the online supplement.

### 2.3 Statistical analysis

Stata, version 16.1 was used for statistical analyses^28^. Samples with missing primary outcome data or exposome variables were excluded from the study, and those with missing covariates were excluded from related analyses. The numbers of missing variables are reported in Table S8 in the online supplement. Similar to prior studies^12,29^, PRS-SCZ and ES-SCZ were dichotomized at the 75th percentile (hereafter PRS-SCZ_75_ and ES-SCZ_75_, respectively).

#### 2.3.1 Main Analyses

The main effects of PRS_cs-auto_-SCZ_75_ and ES-SCZ_75_ at 2-year follow-up on distressing PE at 3-year follow-up were tested in separate models using multilevel logistic regression analysis with random intercepts for collection sites and family. Past-month distressing PEs at 3-year follow-up were used as the primary outcome to test the prospective association of the ES-SCZ adjusted for distressing PEs from prior waves (i.e., baseline to 2-year follow-up). Lifetime distressing PEs, repeating distressing PEs ≥ 2 waves, repeating distressing PEs ≥ 3 waves, and persisting distressing PEs were used as secondary outcomes to explore the influences of PRS-SCZ and ES-SCZ on varying degrees of persistence of distressing PEs. Consistent with previous work^12,29^, additive models were used to test the interaction effects between PRS_cs-auto_-SCZ_75_ and ES-SCZ_75_. As an indicator of the departure from additivity, relative excess risk due to interaction (RERI)^30^ was calculated through delta method^31^ using the Stata “nlcom” postestimation command. RERI greater than zero reflects additional risk over the summation of independent risks of genetic and environmental risk status.

Similar to prior studies^11,32^, the models were adjusted for two sets of covariates: Model 1) age and sex; Model 2) age, sex, family income, and parental education. Analyses including PRS_cs-auto_-SCZ_75_ were additionally adjusted for the first ten ancestrally informative genetic principal components. The nominal significance threshold was set to *p* <0.05 for the primary outcome. As the secondary outcomes were of explorative nature, we did not apply a correction for multiple testing^33^.

#### 2.3.2 Sensitivity Analyses

As PRS-cs-auto is a relatively new method, PRS_ice_-SCZ75 generated by the clumping and thresholding method was used to test the main genetic effect and its interaction with the environment on distressing PEs to ensure the comparability with existing studies. The main effects of ES-SCZ_75_ on distressing PEs were also tested in the whole ABCD study sample as a sensitivity analysis.

## 3. Results

### 3.1 Sample characteristics

Table 1 shows baseline sociodemographic characteristics of the sample included in the main analyses (n=5,122) and exposure frequencies at 2-year follow-up. At 3-year follow-up, participants were 12.9 years on average (SD=0.65, range 11.5-14.6 years), with 11.0% (n=561) having distressing PEs in the past month. Lifetime (≥ 1 wave), repeating ≥ 2 waves, repeating ≥ 3 waves, and persisting (all 4 waves) distressing PEs were endorsed by 1,751 (34.2%), 741 (14.5%), 289 (5.6%), and 98 (1.9%) participants, respectively.

**Table 1.** Sample characteristics at baseline and lifetime exposure up to 2-year follow-up.

| Characteristics at baseline | Mean | SD |
| --- | --- | --- |
| Age (years) | 9.93 | 0.63 |
| Parental educational attainment (years) | 18.2 | 1.7 |
|  | **n** | **%** |
| Male | 2,721 | 53.1 |
| Family income |  |  |
| < \$5,000 | 28 | 0.6 |
| \$5,000-\$11,999 | 34 | 0.7 |
| \$12,000-\$15,999 | 30 | 0.6 |
| \$16,000-\$24,999 | 78 | 1.6 |
| \$25,000-\$34,999 | 127 | 2.6 |
| \$35,000-\$49,999 | 268 | 5.5 |
| \$50,000-\$74,999 | 668 | 13.6 |
| \$75,000-\$99,999 | 841 | 17.1 |
| \$100,000-\$199,999 | 2,058 | 41.9 |
| ≥ \$200,000 | 780 | 15.9 |
| **Lifetime exposure**<br>**up to 2-year follow-up** | **n** | **%** |
| Physical abuse | 38 | 0.7 |
| Emotional abuse | 52 | 1.0 |
| Sexual abuse | 126 | 2.5 |
| Physical neglect | 962 | 18.8 |
| Emotional neglect | 118 | 2.3 |
| Bullying | 1,486 | 29.0 |
| Winter birth | 1,571 | 30.7 |
| Hearing impairment | 365 | 7.1 |
| Cannabis use | 8 | 0.2 |

### 3.2 Main effects of PRS-SCZ

Primary analysis revealed that PRS_cs-auto_-SCZ_75_ was not significantly associated with past-month distressing PEs at 3-year follow-up (Table 2). However, secondary analyses showed that PRS_cs-auto_-SCZ_75_ was significantly associated with lifetime distressing PEs (Model 1: OR 1.29 [95% CI 1.08, 1.53], *p*=.004; Model 2: OR 1.22 [95% CI 1.03, 1.45], *p*=.022), and repeating distressing PEs ≥ 2 waves (Model 1: OR 1.34 [95% CI 1.08, 1.65], *p*=.007; Model 2: OR 1.28 [95% CI 1.03, 1.58], *p* =.025), but not repeating distressing PEs ≥ 3 waves and persisting distressing PEs at 3-year follow-up (Table 2). Sensitivity analysis confirmed significant associations of PRS_ice_-SCZ_75_ with lifetime distressing PEs only when adjusted for age and sex (OR 1.22 [95% CI 1.02, 1.45], *p*=.025) but not when additionally adjusted for family income and parental education (OR 1.18 [95% CI 0.99, 1.40], *p*=.066). No other significant association in the main analyses was confirmed (Table S9 in the online supplement).

**Table 2.** Main associations of PRS_cs-auto_-SCZ_75_ with distressing PEs at 3-year follow-up

| Distressing PEs | Model 1 (N = 5,122) |  |  | Model 2 (N = 4,912) |  |  |
| --- | --- | --- | --- | --- | --- | --- |
|  | OR | 95% CI | *p*-value | OR | 95% CI | *p*-value |
| Past-month | 1.22 | 0.98 to 1.50 | .072 | 1.18 | 0.95 to 1.47 | .143 |
| Lifetime ($\geq 1$ wave) | 1.29 | 1.08 to 1.53 | **.004** | 1.22 | 1.03 to 1.45 | **.022** |
| Repeating ($\geq 2$ waves) | 1.34 | 1.08 to 1.65 | **.007** | 1.28 | 1.03 to 1.58 | **.025** |
| Repeating ($\geq 3$ waves) | 1.19 | 0.87 to 1.62 | .269 | 1.15 | 0.83 to 1.59 | .397 |
| Persisting (all 4 waves) | 0.94 | 0.56 to 1.56 | .799 | 0.84 | 0.49 to 1.45 | .534 |

Model 1: Adjusted for age and sex; Model 2: Adjusted for age, sex, family income, and parental education.
*p*-values <.05 were bolded. PEs, psychotic experiences; OR, odds ratio; CI, confidence interval.

### 3.3 Main effects of ES-SCZ

ES-SCZ_75_ at 2-year follow-up was significantly associated with past-month distressing PEs (Model 1: OR 1.45 [95% CI 1.19, 1.75], *p* <.001; Model 2: OR 1.37 [95% CI 1.12, 1.68], *p* =.002), as well as all secondary outcomes at 3-year follow-up: lifetime distressing PEs (Model 1: OR 2.77 [95% CI 2.31, 3.31], *p* <.001; Model 2: OR 2.55 [95% CI 2.13, 3.05], *p* <.001), repeating distressing PEs ≥ 2 waves (Model 1: OR 3.16 [95% CI 2.54, 3.93], *p* <.001; Model 2: OR 2.93 [95% CI 2.35, 3.65], *p* <.001), repeating distressing PEs ≥ 3 waves (Model 1: OR 3.93 [95% CI 2.86, 5.40], *p* <.001; Model 2: OR 3.47 [95% CI 2.51, 4.78], *p* <.001), and persisting distressing PEs (Model 1: OR 3.65 [95% CI 2.34, 5.70], *p* <.001; Model 2: OR 3.22 [95% CI 2.02, 5.13], *p* <.001) (Table 3). As shown in Figure 1, the effects of ES-SCZ_75_ on distressing PEs tend to be stronger when the experiences become more persistent. Sensitivity analysis of the whole sample confirmed all significant associations (Table S10 in the online supplement).

**Table 3.**
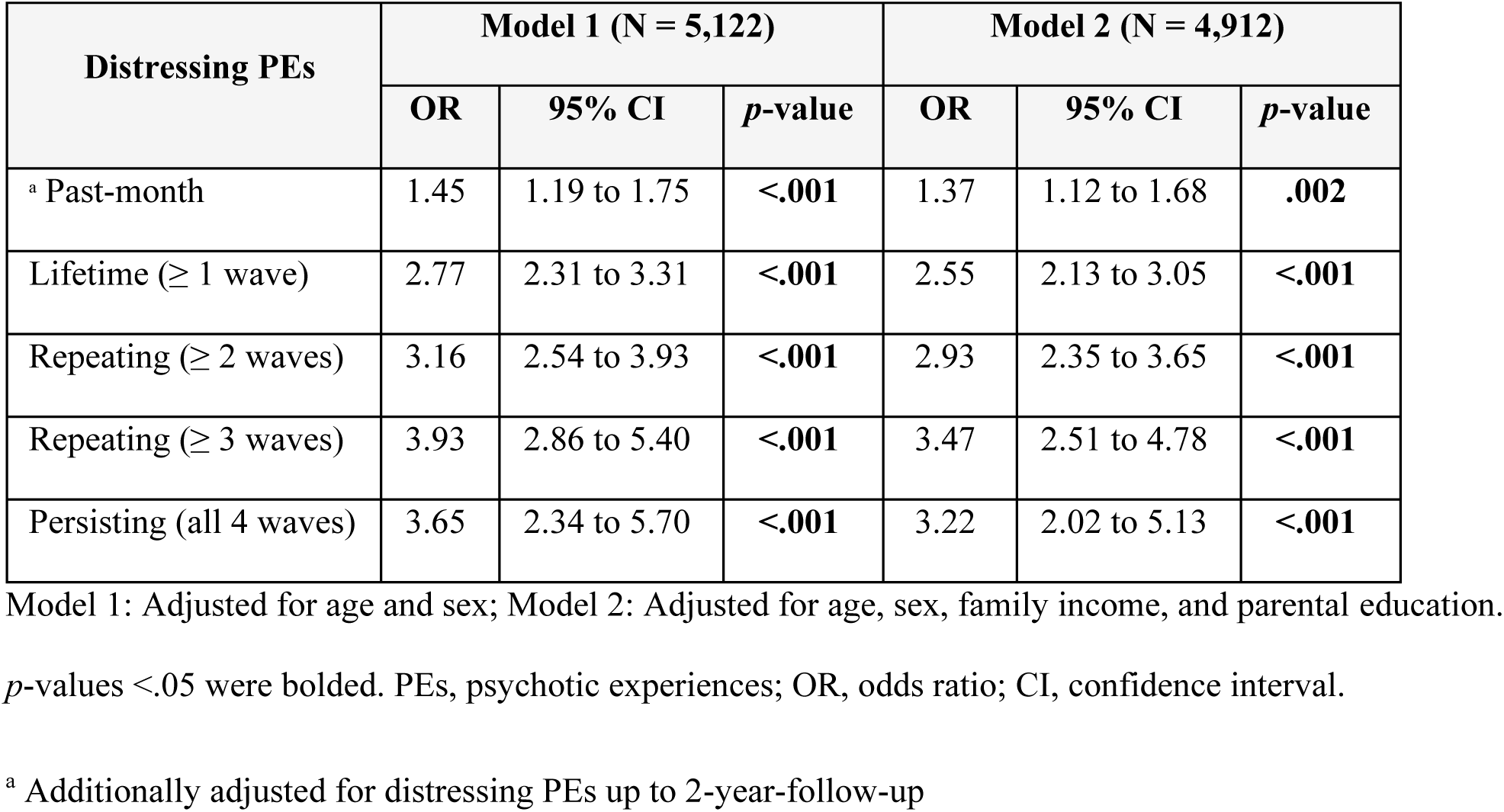
Main associations of ES-SCZ_75_ with distressing PEs at 3-year follow-up

**Figure 1.**
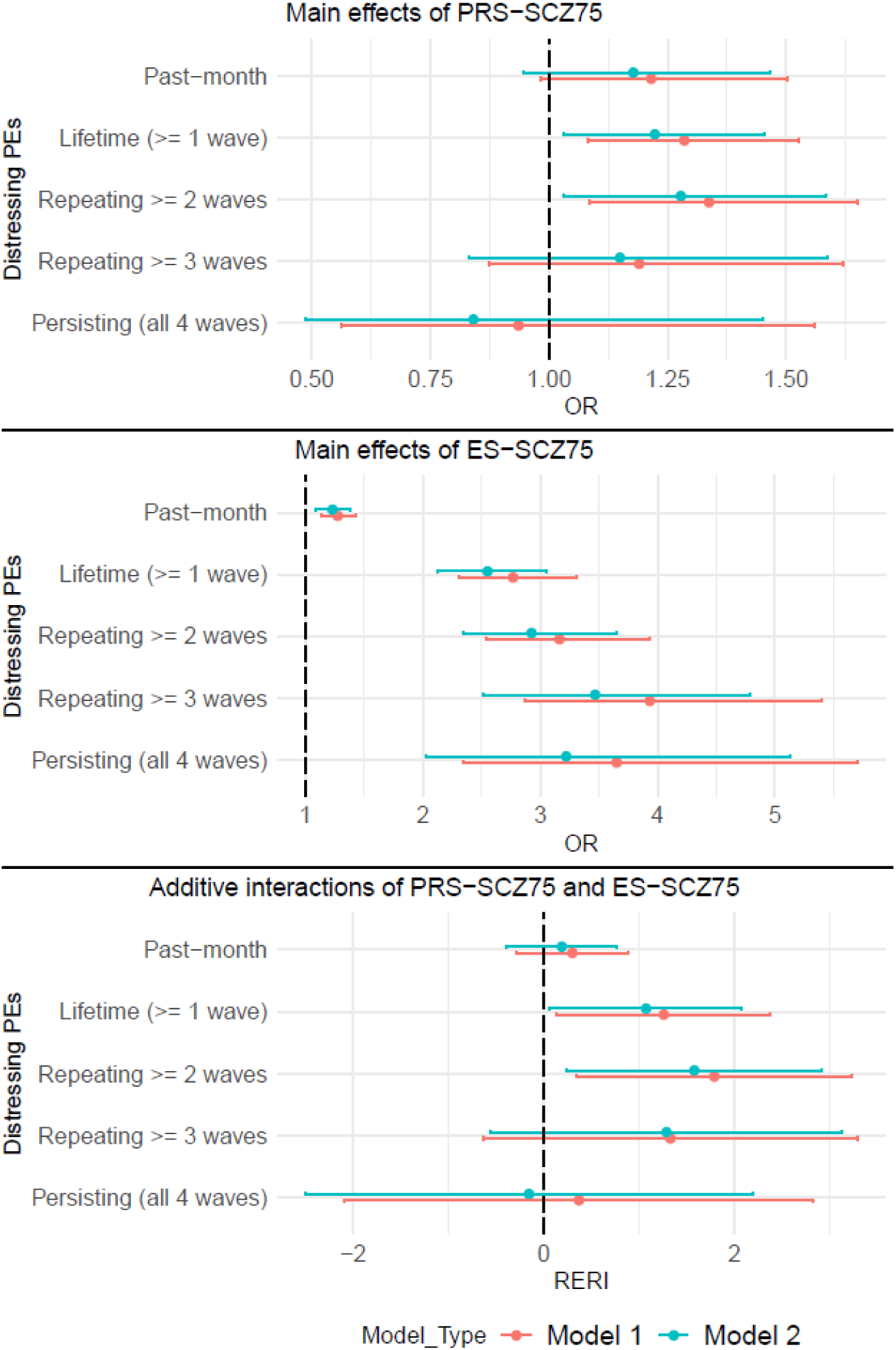
Visualization of main and interacting associations of PRS_cs-auto_-SCZ_75_ and ES-SCZ_75_ at 2-year follow-up on distressing PEs with varying degrees of symptom persistence at 3-year follow-up. Model 1: adjusted for age and sex; Model 2: adjusted for age, sex, family income, and parental education; Upper panel: Main effects of PRS_cs-auto_-SCZ_75_ (additionally adjusted for 10 PCs); Middle panel: Main effects of ES-SCZ_75_ (additionally adjusted for distressing PEs up to 2-year follow-up for past-month distressing PEs); Lower panel: Additive interaction between PRS_cs-auto_-SCZ_75_ and ES-SCZ_75_ (additionally adjusted for 10 PCs for all outcomes and distressing PEs up to 2-year follow-up for past-month distressing PEs). OR, odds ratio; RERI, relative excess risk due to interaction.

### 3.4 Additive interaction

Primary analysis did not find a significant additive interaction between PRS_cs-auto_-SCZ_75_ and ES-SCZ_75_ at 2-year follow-up in predicting past-month distressing PEs at 3-year follow-up (Table S11 in the online supplement & Figure 2A). For secondary outcomes, significant positive additive interactions were found for lifetime distressing PEs (Model 1: RERI 1.26 [95% CI 0.14, 2.38], *p* =.027; Model 2: RERI 1.07 [95% CI 0.06, 2.08], *p*=.037) (Figure 1A), and repeating distressing PEs ≥ 2 waves (Model 1: RERI 1.79 [95% CI 0.35, 3.23], *p* =.015; Model 2: RERI 1.58 [95% CI 0.24, 2.92], *p*=.021) (Figure 2B), but not repeating distressing PEs ≥ 3 waves and persisting distressing PEs at 3-year follow-up (Table S11 in the online supplement). Sensitivity analyses using PRS_ice_-SCZ_75_ did not confirm the significant interactions (Table S12 in the online supplement).

**Figure 2.**
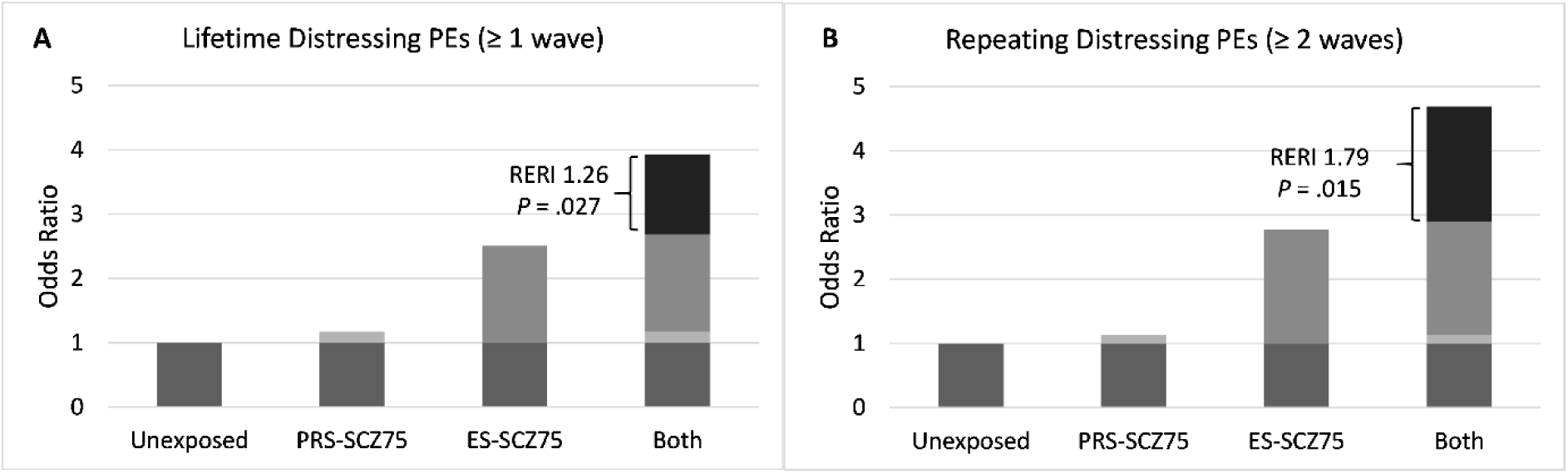
Additive interaction between PRS_cs-auto_-SCZ_75_ and ES-SCZ_75_ on lifetime distressing PEs (A) and repeating distressing PEs ≥ 2 waves (B), adjusted for age, sex, 10 PCs (Model 1). OR, odds ratio; RERI, relative excess risk due to interaction.

## 4. Discussion

To our knowledge, this is the first longitudinal investigation of the independent and joint associations of genomic and exposomic liabilities for schizophrenia with distressing PEs and their persistence in early adolescence. Our findings show that PRS-SCZ was associated with distressing PEs and additively interacted with ES-SCZ in predicting lifetime distressing PEs reported at least in one or two of four annual assessments. We also found a significant association between ES-SCZ in the previous year and distressing PEs reported in the following year. Our secondary analyses revealed that the strength of associations of ES-SCZ with distressing PEs increased as a function of the persistence of the experiences.

A significant association between PRS-SCZ and distressing PEs confirms the notion that PEs share genetic liability with schizophrenia across the psychosis continuum^6^. This aligns with prior studies of PRS-SCZ and subclinical psychosis expression in late adolescence^9,10^ and adulthood^12^ and concords with a cross-sectional analysis of the ABCD dataset at baseline^11^. However, we did not find an association between PRS-SCZ and distressing PEs that repeated for three or more waves. It might be likely that the lower number of participants with more persistent distressing PEs might not have enough statistical power to detect the association.

Similarly, previous studies reporting a non-significant association between PRS-SCZ and PEs also appeared to suffer from issues that might reduce statistical power: relatively small sample sizes (<2,500), less-powered discovery GWAS, and mixed ethnic population^7,14^. Considering that the genetic overlap between SCZ and PEs was larger in adults compared to adolescents^34^ and the prevalence of repeating distressing PEs would be growing over time, a replication study when the ABCD cohort reaches early adulthood might provide a deeper insight into the age-dependent relation between genetics and symptom persistence.

Compared to PRS-SCZ, ES-SCZ appeared to be more strongly associated with distressing PEs across all outcomes and models. This finding conforms with previous research showing that environmental risk factors for schizophrenia explain more variance of subthreshold positive symptoms compared to PRS-SCZ^35^.

Furthermore, we demonstrated that the magnitude of the association of ES-SCZ with distressing PEs increased as a function of persistence. This agrees with the prior analysis of two large prospective general population cohorts, showing that the persistence rate of PEs is progressively increased with the increasing environmental risk exposures^36^. As persistence of distressing PEs is considered a herald of not only future psychotic disorders but also other psychopathology^2,5^, our finding implies that modifying environmental risk exposures could be a promising strategy to prevent psychopathology in youth.

Apart from the independent effects of PRS-SCZ and ES-SCZ on distressing PEs, we explored their joint effects and found significant positive additive interactions between both risk factors on distressing PEs that emerged at least once or twice during ages 9 to 13. However, we did not find significant interactions in the sensitivity analyses using the PRS-ice, which might reflect a lower performance of PRS-ice compared with PRS-cs-auto^22^. Agreeing with our main findings, a previous study showed that PRS-SCZ and ES-SCZ additively interacted in predicting schizotypal traits among healthy adult participants and non-affected siblings of patients with schizophrenia^12^. Another study using large adolescent twin cohorts from the UK and Sweden demonstrated gene-by-environment interaction for subclinical psychosis expression in late adolescence^37^. On the contrary, a research of a Brazilian children cohort demonstrated neither main nor interacting effects of polygenic and poly-environmental risk scores for schizophrenia on PEs and speculated that these null associations might be partly explained by the population admixture of their sample^14^. Although more studies using large cohorts and improved predictive performance of multi-ancestry PRS are required, our findings, together with early evidence, support the synergism between genetic and exposomic risks for schizophrenia in determining subthreshold psychosis expression.

Prior research from the ABCD study has shown that children with distressing PEs presenting at least twice across three waves exhibited the greatest impairment compared to those with transient and/or non-distressing PEs^4^. When combined with such evidence, our findings corroborate the psychosis proneness-persistence-impairment model^38^. Guided by meta-analytical evidence, the model suggests that, despite the transient nature of most PEs, these PEs could abnormally persist and lead to subsequent impairment if the person is additionally exposed to environmental exposures, which also interact with the individual genetic background^38^. Indeed, empirical evidence shows that persistent trajectory of PEs is associated with secondary distress and strongly predicts a transition from subclinical to clinical psychosis longitudinally^39^. Considering that the prevalence of PEs and their persistence peaks in adolescence^5^, screening for psychosis proneness with subsequent early intervention aiming at reducing environmental risk exposure could be an effective strategy to prevent the persistence of distressing PEs with subsequent impairment and eventual development of clinical psychosis.

Despite all the above findings, our research should be considered in light of some limitations. First, PEs were assessed using a self-reported questionnaire that could be subject to recall bias^5^. However, the validity of a self-screening questionnaire for PEs has been tested against clinical interviews in adolescents with acceptable sensitivity and specificity^40^. More specific to the ABCD study, Karcher et al.^41^ showed that the association between parent-rated (through Child Behavior Checklist – psychosis) and child-rated (using PQ-BC) PEs is poor, with the latter being more associated with known psychosis risk factors than the former, supporting the validity of the child’s report. Nonetheless, the one-month timeframe inquired by PQ-BC might limit the detection of PEs that fall outside the assessment window, potentially resulting in an underestimation of the association.

Second, to ensure a good performance of PRS-SCZ derived from the most updated GWAS of the same ancestries^21,42^, we restricted our analyses to European subpopulations. This approach might limit the generalizability of our findings to non-European ancestries and decrease the statistical power of analyses for less prevalent outcomes or small effect sizes. Indeed, we failed to detect the main effects of PRS-SCZ and its interaction with ES-SCZ for more persistent distressing PEs, even though the larger main effects of ES-SCZ on distressing PEs could be confirmed at all degrees of persistence across adjusted models and sensitivity analyses. Therefore, a replication analysis using a large non-European cohort is required to improve the generalizability of the findings and ensure adequate power to detect potentially smaller influences of genetic factors and their interaction with environmental exposure in predicting the persistence of distressing PEs.

Lastly, although the predictive capacity of ES-SCZ has been well-tested in both clinical and non-clinical populations^12,18^, increasing availability of comprehensive data resources linked to electronic health records and geolocated data in the future could allow for improving ES-SCZ with additional exposures. Moreover, integrating multiple sources of information to more accurately determine the risk exposure would complement our findings, which mostly relied on parent and child reports.

In conclusion, our findings highlight the shared genomic and exposomic etiology between subthreshold PEs and clinical psychosis expression, as well as their interaction underlying the persistence of distressing PEs. Although our findings require further replication, public health policy reforms aiming at minimizing environmental risk in young adolescents, particularly in those with increased genetic liability for schizophrenia, could be a promising strategy to improve population mental well-being by preventing the progression of subclinical psychosis to clinically significant psychopathology.

## Data Availability

All data produced in the present study are available upon reasonable request to the authors.

## Acknowledgements

This work was supported by the National Institute of Health (Grant No. U01DA041120 [to DMB]; Grant Nos. K23MH121792 and L30MH120574 [to NRK]; Grant Nos. MH109532 and DA032573 [to AA]; Grant No. F32AA027435 [to ECJ]; Grant No. T32-DA007261 [to ASH]; and Grant Nos. R01-AG045231, R01-HD083614, R01-AG052564, R21-AA027827, and R01-DA046224 [to RB]).

Data used in the preparation of this article were obtained from the Adolescent Brain Cognitive Development (ABCD) Study (https://abcdstudy.org), held in the National Institute of Mental Health Data Archive. This is a multisite, longitudinal study designed to recruit more than 11,500 children age 9–10 years and follow them over 10 years into early adulthood. The ABCD Study is supported by the National Institutes of Health and additional federal partners under Grant Nos. U01DA041022, U01DA041025, U01DA041028, U01DA041048, U01DA041089, U01DA041093, U01DA041106, U01DA041117, U01DA041120, U01DA041134, U01DA041148, U01DA041156, U01DA041174, U24DA041123, and U24DA041147. A full list of supporters is available at https://abcdstudy.org/nih-collaborators. A listing of participating sites and a complete listing of the study investigators can be found at https://abcdstudy.org/principalinvestigators.html. ABCD consortium investigators designed and implemented the study and/or provided data but did not necessarily participate in analysis or writing of this report. This manuscript reflects the views of the authors and may not reflect the opinions or views of the National Institutes of Health or ABCD consortium investigators.

The ABCD data repository grows and changes over time. The ABCD data used in this report came from http://dx.doi.org/10.15154/1523041 (data release 4.0) and http://dx.doi.org/10.15154/z563-zd24 (data release 5.1)

## Author Contributions

M.D.V and T.P. contributed equally to this work (co-first authorship). A.F. and S.G. contributed equally to this work (co-last authorship). S.G., T.P., L.K.P., and M.D.V. developed the study design. Under the supervision of S.G., M.D.V., and T.P. processed the data, and B.D.L, A.A.M., and T.P. performed the analyses. S.G., G.S., and A.F. supervised the process. J.V.O, B.P.F.R., and S.G. acquired funding. T.P. and M.D.V. prepared the first draft of the manuscript. S.G. provided critical revision of the manuscript for important intellectual content. R.B. reviewed the manuscript and provided comments/suggestions. All authors contributed to the final version and have approved the final manuscript.

