## Supplementary material for "Independent and joint effects of genomic and exposomic loads for schizophrenia on distressing and persisting psychotic experiences in adolescence"

1. Quality Control, Imputation and Polygenic Risk Score calculations - ABCD Genotype Data Release 5.1……………………………………………………….p. 2

2. ABCD variables used for calculating ES-SCZ and ABCD variables used as covariates…………………………………………………......................p. 13

3. Tables of data missingness………………………………..…………….…p. 25

4. Tables of results ……………………..……………………………………p. 26

### Quality Control, Imputation and Polygenic Risk Score calculations - ABCD Genotype Data Release 5.1

The ABCD sample consisted in 11,101 individuals from which genomic DNA was extracted from buccal mucosa using Chemagic STAR DNA Saliva 4k Kit (Hamilton Robotics, Reno, NV, United States). Rutger University Cell and DNA Repository (RUCDR) performed a genome-wide genotyping using the Affymetrix NIDA SmokeScreen Array (Affymetrix, Santa Clara, CA, USA), resulting in the genotyping of 733,329 Single-Nucleotide Polymorphisms (SNPs). The genotyped Quality Controlled (QCed) data was in binary PLINK format and contained 516,598 genetic variants referenced in positive strand and aligned to GRCh37 (hg19). Further details can be found in the NDA 4.0 Release Notes – Genetics (https://nda.nih.gov/study.html?id=1299).

#### 1. Pre-imputation Quality Control

Pre-imputation Quality Control (QC) was performed using PLINK 1.9[1]. The initial phase involved genotypic QC, which proceeded as follows: i. RUCDR performed DNA quality controls based on calling signals and variant call rates. ii. ABCD DAIRC performed the subsequent study-based QC process, following the recommendation of Ricopili pipeline (https://nda.nih.gov/study.html?id=1299).

Following the genotype QC, a strict SNP QC was performed. This process was divided into two phases: SNP QC and sample QC. The SNP QC consisted of the following steps: (1) removing SNPs with a Minor Allele Frequency (MAF) < 10% (2) and a Hardy-Weinberg Equilibrium p-value < 1e-5, (3) excluding SNPs located in long Linkage Disequilibrium (LD) regions, (4) removing non-autosomal SNPs, (5) pruning SNPs on maximum LD $r^2 = 0.5$. The Sample QC consisted of (6) removing bad QC samples, (7) removing samples which were mismatched between genetic sex and reported sex, (8) removing samples with problematic genotyping batch, and (9) heterozygosity outliers (+/- 2 Standard Deviation (SD) samples. The detail of QC steps is shown in Table S1.

**Table S1** - Pre-imputation QC steps.

| Step | SNP start | SNP end | Subjects start | Subjects end |
|---|---|---|---|---|
| Strict SNP QC for removal of bad samples – remove SNPS with MAF < 10% and HWE 1e-5 | 516,598 | 230,733 | 11,101 | 11,101 |
| Strict SNP QC – Remove SNPs in long LD regions | 230,733 | 223,421 | 11,101 | 11,101 |
| Strict SNP QC – Remove insertions and deletions | 223,421 | 222,422 | 11,101 | 11,101 |
| Strict SNP QC – Only autosomes (Chr 1 – 22) | 222,422 | 216,956 | 11,101 | 11,101 |
| Strict SNP QC – Prune SNPs on 0.5 max LD | 216,956 | 141,489 | 11,101 | 11,101 |
| Sample QC – Remove bad QC samples | 141,489 | 141,489 | 11,101 | 11,099 |
| Sample QC – Remove mismatch sex pheno samples | 141,489 | 141,489 | 11,099 | 11,061 |
| Sample QC – Remove bad batch samples | 141,489 | 141,489 | 11,061 | 10,979 |
| Sample QC – Remove samples as heterozygosity outliers (+/- 2SD) | 141,489 | 141,489 | 10,979 | 10,232 |

Note: The steps coloured in blue are the QC steps to generate a strict quality control SNP list, which only used to process bad samples exclusion and to calculate PCs.

#### 1.1 Relatedness checking

From the samples that passed the QC (n = 10,232), a relatedness check was performed (using Identity by descent (IBD) information from plink --genome function). As we can see in Table S2, kinship and relationship of the individuals was checked from the Family ID obtained from the "ABCD ACS Post Stratification Weights" dataset. As shown in Figure 1, the distribution of pi-hat value (Proportion IBD, i.e. P(IBD=2) + 0.5*P(IBD=1)) is within expectation: MZ ~ 1, DZ ~ 0.5 and siblings ~ 0.5. Hence, we did not exclude any samples from this step.

**Table S2** - Relatedness checking. The number of singletons, siblings, twins and triplets in the sample that passed the QC are reported.

| Relatedness | N |
|---|---|
| Singletons | 6,738 |
| Siblings | 1,548 |
| Twins | 1,917 |
| Triplets | 29 |

**Figure S1** - Violin plot describing $\hat{\pi}$ (pi-hat) value distribution among related individuals.

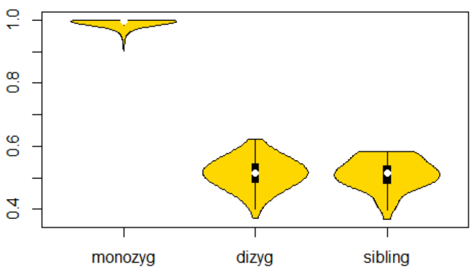


The X axis represents the types of relatedness, categorizing into monozygotic twins (MZ), dizygotic twins (DZ) and siblings. The Y axis represents $\hat{\pi}$ value: (Proportion IBD, i.e. P(IBD=2) + 0.5*P(IBD=1)).

#### 1.2. Principal Component Analysis

A Principal Component Analysis (PCA) was performed on a pruned genetic dataset comprising 141,489 LD-independent SNPs, and 20 Principal Components (PCs) were extracted using the plink --pca function.

First of all, in order to gain insights into the distribution of various populations (AFR = Africans, AMR = Admixed Americans, EUR = Europeans, and EAS = East Asians) within the 1000

Genomes Phase 3 Project[2], PCA plots were conducted. As shown in Figure S2 and Figure S3, the area in which the ancestry of each cohort is defined (EUR and AMR) was circled. This area was determined by calculating the mean +/- 3SD of the first two PCs.

Finally, in order to define only those individuals with European ancestry the ABCD and 1000 genomes datasets were merged. As observed in Figures S4 and S5, those individuals within 1000 genomes EUR area were considered to have European ancestry. The number of individuals and % of the total sample are shown in Table S3.

**Figure S2** – PCA plot of the first 2 PCs of 1000 Genome Phase 3 samples calculated by PLINK 1.9.

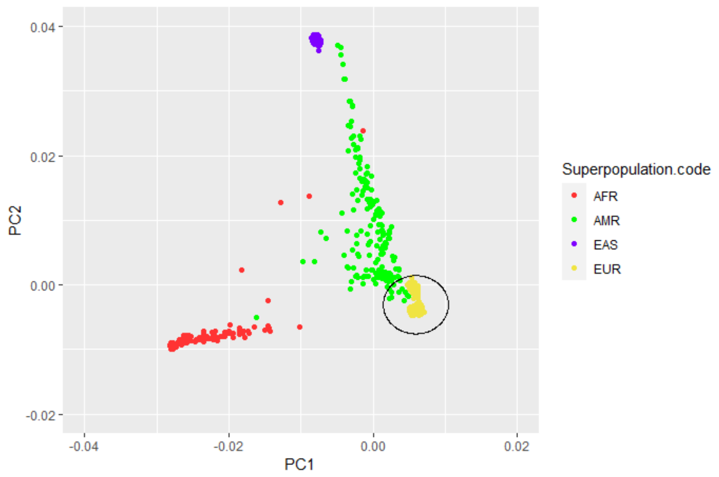


The X and Y axes represent PC1 and PC2 values, respectively. Each population is highlighted in different colours: AFR (Africans, in red, n = 246), AMR (Admixed Americans, in green, n = 181), EAS (East Asians, in purple, n = 286), and EUR (Europeans, in yellow, n = 379). The circled area presents samples defined with European ancestry (mean +/- 3SD for first 2 PCs).

**Figure S3** – PCA plot of the first 2 PCs of 1000 Genome Phase 3 samples calculated by PLINK 1.9.

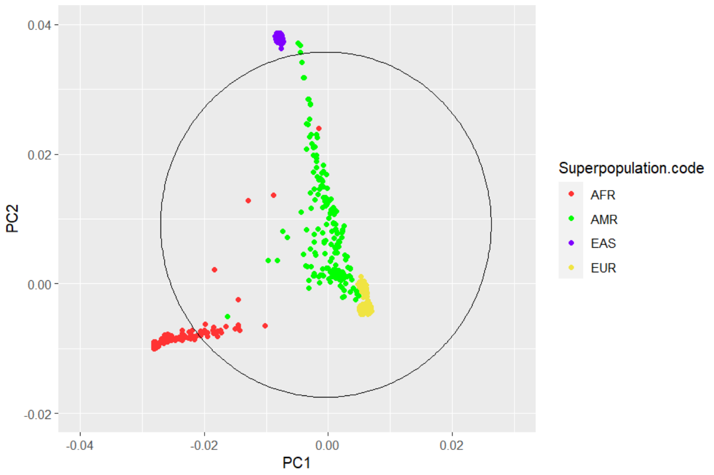


The X and Y axes represent PC1 and PC2 values, respectively. Each population is coloured, and the circled area presents samples with Admixed American ancestry (mean +/- 3SD for first 2 PCs).

**Figure S4 –** PCA plot of the first 2 PCs of ABCD samples.

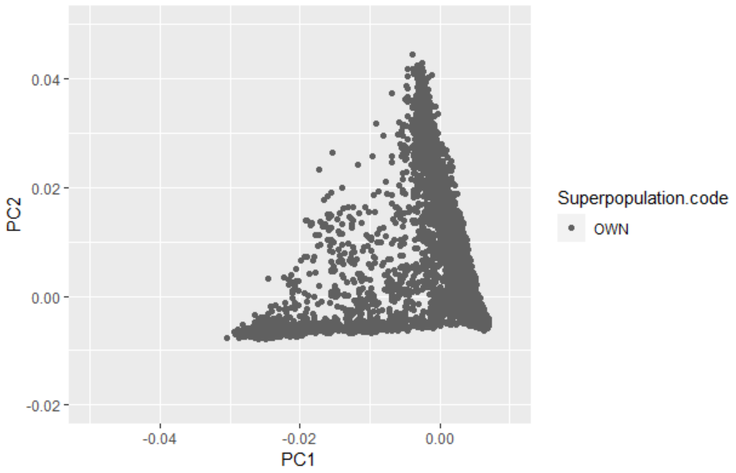


The X and Y axes represent PC1 and PC2 values, respectively. “OWN” cohort is represented (ABCD sample, in grey, n = 10,232).

**Figure S5** – First 2 PCs' PCA plot of the merged ABCD and 1000 Genomes Phase 3 datasets.

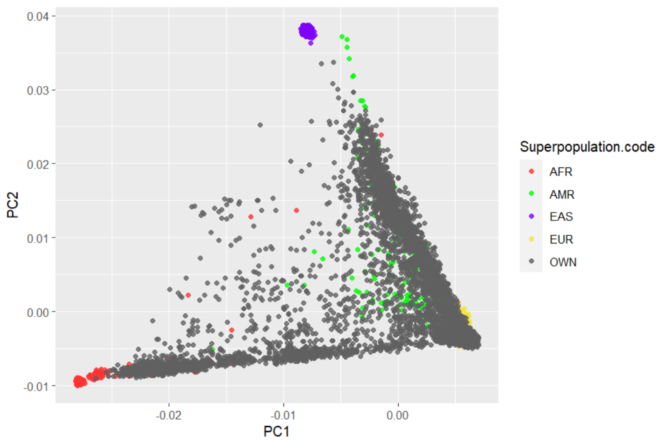


The X and Y axes represent PC1 and PC2 values, respectively. Each population is coloured, and the OWN cohort is represented (ABCD sample, in grey, n = 10,232)

**Table S3** - Distribution of the ABCD sample per ancestry (number of individuals and % from the total sample)

| Sample Ancestry | | |
|---|---|---|
| | N | % |
| Total | 10,232 | 100 |
| EUR | 5,656 | 55.28 |
| AMR | 8,352 | 81.63 |
| AFR | 1,195 | 11.68 |

#### 2. Imputation and Post-imputation Quality Control

Genotypes where phased (Eagle v.2.4) and imputed to the TOPMed (Version R2 on GRCh38; https://imputation.biodatacatalyst.nhlbi.nih.gov) reference panel using MiniMac 4 via the TOPMed Imputation Server,[3] finally yielding 292,136,462 SNPs. Subsequently, the resulting VCF files were converted into dosage PLINK files and then to hard-call genotypes format.

A post-imputation SNP QC was performed using PLINK 1.9[1] according to the following criteria: (1) removed SNPs with a MAF < 1%, (2) Imputation Quality Information (INFO) < 0.9, (3) ambiguous and multiallelic SNPs, (4) insertions and deletions variants, and finally (5) SNPs with HWE p-value < 1e-6. The detail of QC steps is shown in Table S4. In the end, there are 10,232 subjects and 5,909,115 SNPs included in the imputed ABCD data.

**Table S4** - Post-imputation QC describing the SNPs and subjects excluded in all of the steps.

| **SNP QC Step** | **SNP start** | **SNP end** | **Subjects start** | **Subjects end** |
|---|---|---|---|---|
| Remove SNPs with MAF < 1% | 292,136,462 | 11,635,718 | 10,232 | 10,232 |
| Remove SNPs with INFO < 0.9 | 11,635,718 | 9,999,328 | 10,232 | 10,232 |
| Remove ambiguous SNPs | 9,999,328 | 8,551,596 | 10,232 | 10,232 |
| Remove multiallelic SNPs | 8,551,596 | 8,224,505 | 10,232 | 10,232 |
| Remove insertions and deletions | 8,224,505 | 7,659,218 | 10,232 | 10,232 |
| Remove SNPs with HWE 1e-6 | 7,659,218 | 5,909,115 | 10,232 | 10,232 |

#### 3. PRS calculation

For target dataset, there were 10,232 subjects and 5,909,115 imputed SNPs included in ABCD data. PRS scores were based on associated alleles and effect sizes reported in the GWAS summary statistics from the SCZ PGC freeze 3 European subsample[4].

##### 3.1 PRS generation using PRSice-2

The SNPs in the base data (n= 7,659,767) underwent QC measures, resulting in 5,428,447 variants used to calculate PRS. The QC steps involved eliminating multiallelic SNPs, SNPs with bad imputation quality (INFO score below 0.9), ambiguous SNPs, and insertions and deletions variants. Extraction common SNPs from target (ABCD samples), base (GWAS summary statistics) and reference (1000 Genomes Phase 3)[2] datasets, and subsequently excluding SNPs located in 20 complex-LD regions.[5] Following these procedures, the remaining SNPs were 3,878,700.

The remained SNPs were clumped in two rounds using PLINK 1,9.[1]

- Round 1: physical distance threshold 250kb and LD threshold ($R^2$) 0.5
- Round 2: distance 5000kb, LD threshold 0.2

After conducting two rounds of clumping, we were left with 226,418 SNPs for PRS calculations. Subsequently, PRS calculations were performed using PRSice-2[6] for 12 GWAS p-value thresholds: 5 x $10^{-8}$, 5 x $10^{-7}$, 5 x $10^{-6}$, 5 x $10^{-5}$, 5 x $10^{-4}$, 5 x $10^{-3}$, 0.05, 0.1, 0.2, 0.3, 0.4 and 0.5. The detail of PRS steps is shown in Table S5.

**Table S5** - Base Data QC pre- PRS calculation using PRSice2.

| Step | Base Data |
|---|---|
| | SCZ PGC3 (EUR) |
| Initial SNPs | 7,659,767 |
| SNP QC -- Remove multiallelic SNPs | 7,651,642 |
| SNP QC -- Remove SNPs with INFO <= 0.9 | 6,401,110 |
| SNP QC - Remove ambiguous SNPs | 5,428,447 |
| SNP QC -- Remove insertions and deletions | 5,428,447 |
| Common SNPs between Target, Base and Reference Data + Remove LD regions | 3,878,700 |
| Cumming Round 1 (kb = 250, r2 = 0.5) | 535,466 |
| Clumping Round 2 (kb = 5000, r2 = 0.2) | 226,418 |

#### 3.2 PRS generation using PRS-cs-auto

PRS were generated using the PRS-CS (Polygenic Risk Score-Continuous Shrinkage) software package approach.[7] For this process, we used the PRS-CS “auto” function, which employs a fully Bayesian approach. This approach automatically learns the global shrinkage parameter($\phi$) from the available data. Unlike other methods, it does not prune SNPs based on a specific p-value threshold or perform clumping for independent SNPs. Instead, it assumes a general distribution of effect sizes across the genome and considers for LD between SNPs using an external LD reference panel (in our case, the 1000 Genomes Phase 3 European samples panel). The SNPs in the base data (n= 7,659,767) underwent QC measures, resulting in 742,011 variants used to calculate PRS. Further details regarding the steps of PRS generation can be found in Table S6.

**Table S6** - Base Data QC pre-PRS calculation using PRS-cs-auto.

| Step | Base Data |
|---|---|
| | SCZ PGC3 (EUR) |
| Initial SNPs | 7,659,767 |
| SNP QC -- Remove multiallelic SNPs | 7,651,642 |
| SNP QC -- Remove SNPs with INFO <= 0.9 | 6,401,110 |
| SNP QC - Remove ambiguous SNPs | 5,428,447 |
| SNP QC -- Remove insertions and deletions | 5,428,447 |
| Common SNPs between Target, Base and External LD Reference Panel + Remove long LD regions from Target Data | 742,011 |

**Table of abbreviations**

AMR = American Population

Chr = Chromosome

Dizyg = Dizygotic

DZ = Dizygotic

EAS = East Asian Population

GWAS = Genome-Wide Association Study

HWE = Hardy-Weinberg Equilibrium

IBD = Identity By Descendent

INFO = Imputation Quality Information

LD = Linkage-Disequilibrium

MAF = Minor Allele Frequency

Monozyg = Monozygotic

MZ = Monozygotic

OWN = Own dataset population (ABCD Sample)

PC = Principal Component

PCA = Principal Component Analysis

Pheno = Phenotype data

PRS = Polygenic Risk Score

PRS-CS = Polygenic Risk Score-Continuous Shrinkage

QC = Quality Control

SCZ = Schizophrenia

SD = Standard Deviation

SNP = Single Nucleotide Polymorphis

### ABCD variables used for calculating ES-SCZ and ABCD variables used as covariates

Lifetime environmental exposures obtained from the ABCD Study dataset were recoded as binary variables (0 = "absent"; 1 = "present") using baseline, 1-year follow-up, and 2-year follow-up data to generate ES-SCZ[1]. The following information describes the procedure of dichotomization in detail.

*Emotional neglect*. Emotional neglect was derived from five items of the *Child Report of Parent Behavior Inventory* (CRPBI)[2]. Children reported perceived levels of parents/primary caregivers' warmth and closeness across a 3-point Likert scale (1 = "Not like him/her"; 2 = "Somewhat like him/her"; 3 = "A lot like him/her"). As well as in Stinson et al.[3], emotional neglect was regarded as present if children answered "Not like him/her" in at least two out of five items of the CRPBI.

*Physical neglect.* Consistent with previous research[3,4], physical neglect was considered present if any of the conditions below had been experienced by youth. Food shortage was reported in the *Parent Demographics Survey* and in the *Longitudinal Parent Demographics Survey* (0 = "No"; 1 = "Yes"). History of maternal or paternal alcohol use that had caused isolating self from family, causing arguments or being drunk a lot was reported by parents in two items of the *Family History Assessment Part 1* (0 = "No"; 1 = "Yes"). Poor parental supervision was considered positive if "Never" or "Almost never" were answered by youth in any of the three items on the *Parental Monitoring Survey*.

*Emotional abuse.* Following Stinson et al.[3], emotional abuse was considered present if parents answered "Yes" to the item "A family member threatened to kill your child" on the PTSD module of the *Kiddie Schedule for Affective Disorders and Schizophrenia for DSM-5*[5,6].

*Physical abuse*. Consistent with previous research[3], physical abuse was considered endorsed when parents answered "Yes" to any of the following items on the *Kiddie Schedule for Affective Disorders and Schizophrenia for DSM-5*: "Shot, stabbed, or beaten brutally by a grown up in the home" or "Beaten to the point of having bruises by a grown up in the home".

*Sexual abuse.* When parents endorsed any of the following items on the *Kiddie Schedule for Affective Disorders and Schizophrenia for DSM-5*: "A grown up in the home touched your child in their privates, had your child touch their privates, or did other sexual things to your child", "An adult outside your family touched your child in their privates, had your child touch their privates or did other sexual things to your child", or "A peer forced your child to do something sexually", sexual abuse was considered present[3].

*Cannabis use.* Lifetime cannabis use was regarded positive (coded as "1") if any of the following condition were met. Having ever used cannabis at least 10 times was reported by youth on the *ABCD Youth Substance Use Interview* at baseline. Use of cannabis at least 10 times during the previous year was reported by youth on the *ABCD Youth Substance Use Introduction and Patterns* at the yearly follow-up, and on *the ABCD Timeline Follow-back Survey Calendar Scores (TLFB)* across the third and fourth waves of follow-up. Use of cannabis 5 times or more during the previous 6 months was reported on the *ABCD Youth Marijuana Illicit Drug Measures* at the mid-year follow-up. Having ever experienced craving, withdrawal symptoms, or tolerance was reported on the *ABCD Parent Diagnostic Interview for DSM-5 (KSADS) Drug Use Disorder Individual Questions*.

*Winter birth.* Winter birth was derived by leveraging the precise date of the interview at baseline and the child's age in months, both available in the *Demographics Survey*. The variable was coded as positive ("1") if birthday fell in December, January, February, or March.

*Hearing impairment.* This variable was considered endorsed when parents answered "Yes" at least once to the questions: "Has she/he ever been to a doctor for any of these things… Hearing Problem" on the *ABCD Parent Medical History Questionnaire* at baseline, and: "Since we last saw you, has she/he been to a doctor for any of these things? Hearing Problem" on the *ABCD Longitudinal Parent Medical History Questionnaire* at follow-up.

*Bullying.* Bullying was considered endorsed when parents answered "Yes" to the question: "Does your child have any problems with bullying at school or in your neighborhood?" on the *ABCD Parent Diagnostic Interview for DSM-5 Background Items Full* at baseline, and/or to the question: "Since we last saw you, did your child have any problems with bullying at school or in your neighbourhood?" on the *ABCD Longitudinal Parent Diagnostic Interview for DSM-5 Background Items Full (KSAD)* at follow-up.

Similar to the previous literature[7], ES-SCZ was calculated by summing the nine exposures multiplied by their log odds (i.e. weighted risks) for schizophrenia and added by 2 for ease of interpretation as in the following formula: (cannabis use*1.31) + (winter birth*0.03) + (hearing impairment*1.18) + (emotional abuse*0.78) + (physical abuse*-0.39) + (sexual abuse*0.86) + (emotional neglect*0.44) + (physical neglect*0.25) + (bullying*1.35) + 2

Table S7 lists in detail the variables used for calculating ES-SCZ and those included as covariates in Model 1 and Model 2. ABCD dataset nomenclature and option levels are reported for each.

**Table S7.** ABCD variables used for calculating ES-SCZ and ABCD variables used as covariates.

| *ABCD VARIABLES USED FOR CALCULATING ES-SCZ* | | | |
|---|---|---|---|
| **Variable** | **ABCD dataset** | **ABCD Variable name** | **Item content** |
| Emotional neglect | ABCD Children's Report of Parental Behavioral Inventory - ce_y_crpbi | crpbi_parent1_y | First caregiver (caregiver participating in study/completing protocol). Makes me feel better after talking over my worries with him/her<br><br>This variable has the following levels:<br>1 = Not like him/her<br>2 = Somewhat like him/her<br>3 = A lot like him/her |
| Emotional neglect | ABCD Children's Report of Parental Behavioral Inventory - ce_y_crpbi | crpbi_parent2_y | First caregiver (caregiver participating in study/completing protocol). Smiles at me very often.<br><br>This variable has the following levels:<br>1 = Not like him/her<br>2 = Somewhat like him/her<br>3 = A lot like him/her |
| Emotional neglect | ABCD Children's Report of Parental Behavioral Inventory - ce_y_crpbi | crpbi_parent3_y | First caregiver (caregiver participating in study/completing protocol). Is able to make me feel better when I am upset.<br><br>This variable has the following levels:<br>1 = Not like him/her<br>2 = Somewhat like him/her<br>3 = A lot like him/her |
| Emotional neglect | ABCD Children's Report of Parental Behavioral Inventory - ce_y_crpbi | crpbi_parent4_y | First caregiver (caregiver participating in study/completing protocol). Believes in showing his/her love for me.<br><br>This variable has the following levels:<br>1 = Not like him/her<br>2 = Somewhat like him/her<br>3 = A lot like him/her |
| Emotional neglect | ABCD Children's Report of Parental Behavioral Inventory - ce_y_crpbi | crpbi_parent5_y | First caregiver (caregiver participating in study/completing protocol). Is easy to talk to.<br><br>This variable has the following levels:<br>1 = Not like him/her<br>2 = Somewhat like him/her<br>3 = A lot like him/her |

| | | | |
|---|---|---|---|
| Physical neglect | ABCD Parent Demographics Survey - abcd_p_demo | demo_fam_exp1_v2 | In the past 12 months, has there been a time when you and your immediate family experienced any of the following: Needed food but couldn't afford to buy it or couldn't afford to go out to get it?<br>This variable has the following levels:<br>0 = No<br>1 = Yes<br>777 = Refuse to answer |
| Physical neglect | ABCD Longitudinal Parent Demographics Survey - abcd_p_demo | demo_fam_exp1_v2_l | Needed food but couldn't afford to buy it or couldn't afford to go out to get it?<br><br>This variable has the following levels:<br>0 = No<br>1 = Yes<br>777 = Refuse to answer |
| Physical neglect | ABCD Parental Monitoring Survey - ce_y_pm | parent_monitor_q1_y | How often do your parents/guardians know where you are?<br><br>This variable has the following levels:<br>1 = Never<br>2 = Almost Never<br>3 = Sometimes<br>4 = Often<br>5 = Always or Almost Always |
| Physical neglect | ABCD Parental Monitoring Survey - ce_y_pm | parent_monitor_q2_y | How often do your parents know who you are with when you are not at school and away from home?<br><br>This variable has the following levels:<br>1 = Never<br>2 = Almost Never<br>3 = Sometimes<br>4 = Often<br>5 = Always or Almost Always |
| Physical neglect | ABCD Parental Monitoring Survey - ce_y_pm | parent_monitor_q3_y | If you are at home when your parents or guardians are not, how often do you know how to get in touch with them?<br><br>This variable has the following levels:<br>1 = Never<br>2 = Almost Never<br>3 = Sometimes<br>4 = Often<br>5 = Always or Almost Always |
| Physical neglect | ABCD Family History Assessment Part 1 - mh_p_fhx | famhx_4d_p___6 | Has ANY blood relative of your child ever had any problems due to alcohol, such as: Marital separation or divorce; Laid off or |

| | | | |
|---|---|---|---|
| | | | fired from work; Arrests or DUIs; Alcohol harmed their health; In an alcohol treatment program; Suspended or expelled from school 2 or more times; Isolated self from family, caused arguments or were drunk a lot. Biological mother.<br><br>This variable has the following levels:<br>0 = No<br>1 = Yes |
| Physical neglect | ABCD Family History Assessment Part 1 - mh_p_fhx | famhx4a_p___6 | Has ANY blood relative of your child ever had any problems due to alcohol, such as: Marital separation or divorce; Laid off or fired from work; Arrests or DUIs; Alcohol harmed their health; In an alcohol treatment program; Suspended or expelled from school 2 or more times; Isolated self from family, caused arguments or were drunk a lot. Biological father.<br><br>This variable has the following levels:<br>0 = No<br>1 = Yes |
| Emotional abuse | ABCD Parent Diagnostic Interview for DSM-5 (KSADS) Traumatic Events - mh_p_ksads_ptsd | ksads_ptsd_raw_765_p | A family member threatened to kill your child.<br><br>This variable has the following levels:<br>0 = No<br>1 = Yes |
| Physical abuse | ABCD Parent Diagnostic Interview for DSM-5 (KSADS) Traumatic Events - mh_p_ksads_ptsd | ksads_ptsd_raw_762_p | Shot, stabbed, or beaten brutally by a grown up in the home.<br><br>This variable has the following levels:<br>0 = No<br>1 = Yes |
| Physical abuse | ABCD Parent Diagnostic Interview for DSM-5 (KSADS) Traumatic Events - mh_p_ksads_ptsd | ksads_ptsd_raw_763_p | Beaten to the point of having bruises by a grown up in the home.<br><br>This variable has the following levels:<br>0 = No<br>1 = Yes |
| Sexual abuse | ABCD Parent Diagnostic Interview for DSM-5 (KSADS) Traumatic Events - mh_p_ksads_ptsd | ksads_ptsd_raw_767_p | A grown up in the home touched your child in their privates, had your child touch their privates, or did other sexual things to your child.<br><br>This variable has the following levels: |

| | | | |
|---|---|---|---|
| | | | 0 = No<br>1 = Yes |
| Sexual abuse | ABCD Parent Diagnostic Interview for DSM-5 (KSADS) Traumatic Events - mh_p_ksads_ptsd | ksads_ptsd_raw_768_p | An adult outside your family touched your child in their privates, had your child touch their privates or did other sexual things to your child.<br><br>This variable has the following levels:<br>0 = No<br>1 = Yes |
| Sexual abuse | ABCD Parent Diagnostic Interview for DSM-5 (KSADS) Traumatic Events - mh_p_ksads_ptsd | ksads_ptsd_raw_769_p | A peer forced your child to do something sexually.<br><br>This variable has the following levels:<br>0 = No<br>1 = Yes |
| Cannabis use | ABCD Youth Substance Use Interview - su_y_sui | first_mj_1b | You mentioned earlier that you have had a puff of marijuana, weed or pot in the past. I want to ask some more questions about that. How many total times have you used marijuana?<br><br>Frequency |
| Cannabis use | ABCD Youth Substance Use Introduction and Patterns - su_y_sui | first_mj_1b_l | How many total times have you used marijuana since the last time we saw you?<br><br>Frequency |
| Cannabis use | ABCD Timeline Follow-back Survey Calendar Scores (TLFB) - su_y_tlfb | su_tlfb_cal_scr_mj_days_cum | Combined count of total days of any type of marijuana (excluding CBD) use during entire cumulative period between sessions (detailed plus estimated TLFB periods). |
| Cannabis use | ABCD Youth Marijuana Illicit Drug Measures - su_y_can_mapi | mapi_q08_l | Please indicate how many times each has happened to you during the last six months while you were smoking marijuana or as the result of your marijuana use: felt that you needed more marijuana than you used to use in order to get the same effect.<br><br>This variable has the following levels:<br>0 = 0<br>1 = 1-2<br>2 = 3-5<br>3 = 6-10<br>4 = 10+ |
| Cannabis use | ABCD Youth Marijuana Illicit Drug Measures - su_y_can_mapi | mapi_q10_l | Please indicate how many times each has happened to you during the last six months while you were smoking marijuana or as the result of your marijuana use: had withdrawal symptoms, that is, felt |

| | | | |
|---|---|---|---|
| | | | sick because you stopped or cut down on smoking.<br><br>This variable has the following levels:<br>0 = 0<br>1 = 1-2<br>2 = 3-5<br>3 = 6-10<br>4 = 10+ |
| Cannabis use | ABCD Youth Marijuana Illicit Drug Measures - su_y_can_cws | su_cws_15 | Craving/wanting to use marijuana.<br><br>This variable has the following levels:<br>0 = None<br>1 = 1<br>2 = 2<br>3 = 3<br>4 = Some<br>5 = 5<br>6 = 6<br>7 = 7<br>8 = 8<br>9 = A Lot<br>10 = 10 |
| Cannabis use | ABCD Parent Diagnostic Interview for DSM-5 (KSADS) Drug Use Disorder Individual Questions - su_y_ksads_sud | ksads_dud_raw_1413_p | Was there ever a time that your child often craved the following drugs? Marijuana (pot, weed, hash, THC, blunts, synthetic marijuana/K2/Spice, marijuana edibles, concentrates such as dabs or shatter).<br><br>This variable has the following levels:<br>0 = No<br>1 = Yes |
| Cannabis use | ABCD Parent Diagnostic Interview for DSM-5 (KSADS) Drug Use Disorder Individual Questions - su_y_ksads_sud | ksads_dud_raw_1483_p | Was there ever a time that your child had the shakes or other bad symptoms after he or she cut down on their use of the following drugs? Marijuana (pot, weed, hash, THC, blunts, synthetic marijuana/K2/Spice, marijuana edibles, concentrates such as dabs or shatter).<br><br>This variable has the following levels:<br>0 = No<br>1 = Yes |
| Cannabis use | ABCD Parent Diagnostic Interview for DSM-5 (KSADS) Drug Use Disorder Individual Questions - su_y_ksads_sud | ksads_dud_raw_1493_p | Since your child has been using the following drugs, has your child found that he or she needs to use a lot more to get the same high?Marijuana (pot, weed, hash, THC, blunts, synthetic marijuana/K2/Spice, marijuana |

| | | | |
|---|---|---|---|
| | | | edibles, concentrates such as dabs or shatter).<br>This variable has the following levels:<br>0 = No<br>1 = Yes |
| Cannabis use | ABCD Youth Mid Year Phone Interview Substance Use - su_y_mypi | mypi_mj_pst_6mo | Did you smoke marijuana, pot, grass, weed, or ganja 5 or more times in the past 6 months, so from last (name of month 6 months ago) until today?<br><br>This variable has the following levels:<br>1 = Yes<br>0 = No<br>777 = Refuse to answer |
| Cannabis use | ABCD Youth Mid Year Phone Interview Substance Use - su_y_mypi | mypi_mj_oils_pst_6mo | Did you use Marijuana Oils or Concentrates, such as hash oil, BHO, butane, or dabs 5 or more times in the past 6 months, so from last (name of month 6 months ago) until today?<br><br>This variable has the following levels:<br>1 = Yes<br>0 = No<br>777 = Refuse to answer |
| Cannabis use | ABCD Youth Mid Year Phone Interview Substance Use - su_y_mypi | mypi_mj_synth_pst_6mo | Did you use "fake" marijuana or synthetics such as K2 or spice 5 or more times in the past 6 months, so from last (name of month 6 months ago) until today?<br><br>This variable has the following levels:<br>1 = Yes<br>0 = No<br>777 = Refuse to answer |
| Cannabis use | ABCD Youth Mid Year Phone Interview Substance Use - su_y_mypi | mypi_mj_edible_pst_6mo | Did you eat marijuana in any food such as cookies, gummy bears, or brownies 5 or more times in the past 6 months, so from last (name of month 6 months ago) until today?<br><br>This variable has the following levels:<br>1 = Yes<br>0 = No<br>777 = Refuse to answer |
| Cannabis use | ABCD Youth Mid Year Phone Interview Substance Use - su_y_mypi | mypi_mj_tinc_pst_6mo | Did you have a marijuana infused alcohol drink or marijuana tincture 5 or more times in the past 6 months, so from last (name of month 6 months ago) until today? |

| | | | |
|---|---|---|---|
| | | | This variable has the following levels:<br>1 = Yes<br>0 = No<br>777 = Refuse to answer |
| Winter birth date | ABCD Parent Demographics Survey - abcd_y_lt | interview_age | Age in months at the time of the interview/test/sampling/imaging.<br><br>Age is rounded to chronological month. If the research participant is 15-days-old at time of interview, the appropriate value would be 0 months. If the participant is 16-days-old, the value would be 1 month. |
| Winter birth date | ABCD Parent Demographics Survey - abcd_y_lt | interview_date | Date on which the interview/genetic test/sampling/imaging/biospecimen was completed. MM/DD/YYYY. |
| Hearing impairment | ABCD Parent Medical History Questionnaire (MHX) - ph_p_mhx | medhx_2i | Has she/he ever been to a doctor for any of these things… Hearing Problem.<br><br>This variable has the following levels:<br>1 = Yes<br>0 = No |
| Hearing impairment | ABCD Longitudinal Parent Medical History Questionnaire - ph_p_mhx | medhx_2i_l | Since we last saw you, has she/he been to a doctor for any of these things? Hearing Problem.<br><br>This variable has the following levels:<br>1 = Yes<br>0 = No |
| Bullying | ABCD Parent Diagnostic Interview for DSM-5 Background Items Full (KSADS-5) - mh_p_ksads_bg | kbi_p_c_bully | Does your child have any problems with bullying at school or in your neighborhood?<br><br>This variable has the following levels:<br>1 = Yes<br>0 = No |
| Bullying | ABCD Longitudinal Parent Diagnostic Interview for DSM-5 Background Items Full (KSAD) - mh_p_ksads_bg | kbi_p_c_bully_l | Since we last saw you, did your child have any problems with bullying at school or in your neighborhood?<br><br>This variable has the following levels:<br>1 = Yes<br>2 = No<br>777 = Decline to answer |
| ***ABCD VARIABLES USED AS COVIARIATES*** | | | |
| **Variable** | **ABCD dataset** | **ABCD Variable name** | **Item content** |

| | | | |
|---|---|---|---|
| Age in months at the time of PE assessment | ABCD Prodromal Psychosis Scale - abcd_y_lt | interview_age | Age in months at the time of the interview/test/sampling/imaging. Age is rounded to chronological month. If the research participant is 15-days-old at time of interview, the appropriate value would be 0 months. If the participant is 16-days-old, the value would be 1 month. |
| Sex | ABCD Parent Demographics Survey - abcd_p_demo | sex | Sex of subject at birth.<br><br>This variable has the following levels:<br>M = Male; F = Female; O=Other; NR = Not reported |
| Baseline family income | ABCD Parent Demographics Survey - abcd_p_demo | demo_comb_income_v2 | What is your TOTAL COMBINED FAMILY INCOME for the past 12 months? This should include income (before taxes and deductions) from all sources, wages, rent from properties, social security, disability and/or veteran's benefits, unemployment benefits, workman's compensation, help from relative (include child payments and alimony), and so on.<br><br>This variable has the following levels:<br>1 = Less than $5,000;<br>2 = $5,000 through $11,999;<br>3 = $12,000 through $15,999;<br>4 = $16,000 through $24,999;<br>5 = $25,000 through $34,999;<br>6 = $35,000 through $49,999;<br>7 = $50,000 through $74,999;<br>8 = $75,000 through $99,999;<br>9 = $100,000 through $199,999;<br>10 = $200,000 and greater.<br>777 = Refuse to answer<br>999 = Don't know |
| Highest parental education | ABCD Parent Demographics Survey - abcd_p_demo | Maximum value of demo_prnt_ed_v2 and demo_prtnr_ed_v2 | What is the highest grade or level of school you have completed or the highest degree you have received?<br><br>This variable has the following levels:<br>0 = Never attended/Kindergarten only; 1 = 1st grade; 2 = 2nd grade; 3 = 3rd grade; 4 = 4th grade; 5 = 5th grade; 6 = 6th grade; 7 = 7th grade; 8 = 8th grade; 9 = 9th grade; 10 = 10th grade; 11 = 11th grade; 12 = 12th grade; 13 = High school graduate; 14 = GED or equivalent; 15 = Some college; 16 = Associate |

| | | | |
|---|---|---|---|
| | | | degree: Occupational; 17 = Associate degree: Academic Program; 18 = Bachelor's degree (ex. BA); 19 = Master's degree (ex. MA); 20 = Professional School degree (ex. MD); 21 = Doctoral degree (ex. PhD); 777 = Refused to answer |

**Table S8.** The number of missing data in European samples

| Variables | Number of missing data (n=5,650) |
|---|---|
| Primary outcome (at 3-year follow-up) | |
| Psychotic experiences | 522 |
| Lifetime exposure (up to 2-year follow-up) | |
| Physical abuse | 6 |
| Emotional abuse | 6 |
| Sexual abuse | 6 |
| Physical neglect | 0 |
| Emotional neglect | 3 |
| Bullying | 0 |
| Winter birth | 1 |
| Hearing impairment | 1 |
| Cannabis use | 1 |
| Covariates | |
| Age (at 3-year follow-up) | 514 |
| Binary sex | 1 |
| Family income | 251 |
| Parental education | 0 |

**Table S9.** Main effects of $PRS_{ice}$-$SCZ_{75}$ generated by on distressing PEs on distressing PEs at 3-year follow-up in the European subsample

| Distressing PEs | Model 1 (N = 5,122) | | | Model 2 (N = 4,912) | | |
|---|---|---|---|---|---|---|
| | OR | 95% CI | *p*-value | OR | 95% CI | *p*-value |
| Past-month (3-year follow-up) | 1.02 | 0.82 to 1.27 | .835 | 0.99 | 0.79 to 1.27 | .919 |
| Lifetime (≥ 1 wave) | 1.22 | 1.02 to 1.45 | **.025** | 1.18 | 0.99 to 1.40 | .066 |
| Repeating (≥ 2 waves) | 1.18 | 0.96 to 1.46 | .118 | 1.12 | 0.90 to 1.39 | .324 |
| Repeating (≥ 3 waves) | 1.13 | 0.83 to 1.54 | .438 | 1.13 | 0.82 to 1.57 | .448 |
| Persisting (all 4 wave) | 1.05 | 0.64 to 1.74 | .834 | 1.04 | 0.61 to 1.77 | .882 |

Model 1: Adjusted for age and sex; Model 2: Adjusted for age, sex, family income, and parental education. P-values <.05 were bolded.

**Table S10.** Main effects of ES-$SCZ_{75}$ at 2-year follow-up on distressing PEs at 3-year follow-up in the whole ABCD study sample

| Distressing PEs | Model 1 (N = 9,621) | | | Model 2 (N = 8,918) | | |
|---|---|---|---|---|---|---|
| | OR | 95% CI | *p*-value | OR | 95% CI | *p*-value |
| [a] Past-month (3-year follow-up) | 1.36 | 1.19 to 1.56 | **<.001** | 1.31 | 1.13 to 1.51 | **<.001** |
| Lifetime (≥ 1 wave) | 2.71 | 2.38 to 3.10 | **<.001** | 2.60 | 2.27 to 2.97 | **<.001** |
| Repeating (≥ 2 waves) | 2.46 | 2.14 to 2.83 | **<.001** | 2.35 | 2.03 to 2.71 | **<.001** |
| Repeating (≥ 3 waves) | 2.74 | 2.27 to 3.31 | **<.001** | 2.60 | 2.13 to 3.16 | **<.001** |
| Persisting (all 4 wave) | 2.81 | 2.10 to 3.75 | **<.001** | 2.56 | 1.91 to 3.43 | **<.001** |

Model 1: Adjusted for age and sex; Model 2: Adjusted for age, sex, family income, and parental education. P-values <.05 were bolded.

[a] Additionally adjusted for distressing PEs up to 2-year-follow-up

**Table S11.** Additive interaction between $PRS_{cs\text{-}auto}$-$SCZ_{75}$ and ES-$SCZ_{75}$ at 2-year follow-up on distressing PEs at 3-year follow-up in the European subsample

| Distressing PEs | | Model 1 (N = 5,122) | | | Model 2 (N = 4,912) | | |
|---|---|---|---|---|---|---|---|
| | | $PRS_{cs\text{-}auto}$-$SCZ_{75}$ = 0 OR (95% CI) | $PRS_{cs\text{-}auto}$-$SCZ_{75}$ = 1 OR (95% CI) | RERI (95% CI) | $PRS_{cs\text{-}auto}$-$SCZ_{75}$ = 0 OR (95% CI) | $PRS_{cs\text{-}auto}$-$SCZ_{75}$ = 1 OR (95% CI) | RERI (95% CI) |
| [a] Past-month (3-year follow-up) | ES-$SCZ_{75}$ = 0 | 1.0 | 1.02 (0.78 to 1.35) p = .868 | 0.30 (-0.29 to 0.89) p = .312 | 1.0 | 1.04 (0.79 to 1.38) p = .759 | 0.19 (-0.39 to 0.77) p = .522 |
| | ES-$SCZ_{75}$ = 1 | 1.38 (1.10 to 1.73) **p = .005** | 1.71 (1.25 to 2.34) **p = .001** | | 1.33 (1.06 to 1.69) **p = .016** | 1.57 (1.13 to 2.17) **p = .007** | |
| Lifetime (≥ 1 wave) | ES-$SCZ_{75}$ = 0 | 1.0 | 1.17 (0.95 to 1.44) p = .142 | 1.26 (0.14 to 2.38) **p = .027** | 1.0 | 1.11 (0.90 to 1.37) p = .322 | 1.07 (0.06 to 2.08) **p = .037** |
| | ES-$SCZ_{75}$ = 1 | 2.51 (2.06 to 3.05) **p < .001** | 3.93 (2.92 to 5.29) **p < .001** | | 2.31 (1.90 to 2.82) **p < .001** | 3.50 (2.59 to 4.72) **p < .001** | |
| Repeating (≥ 2 waves) | ES-$SCZ_{75}$ = 0 | 1.0 | 1.13 (0.86 to 1.49) p = .391 | 1.79 (0.35 to 3.23) **p = .015** | 1.0 | 1.08 (0.81 to 1.43) p = .589 | 1.58 (0.24 to 2.92) **p = .021** |
| | ES-$SCZ_{75}$ = 1 | 2.77 (2.18 to 3.52) **p < .001** | 4.69 (3.35 to 6.57) **p < .001** | | 2.58 (2.02 to 3.29) **p < .001** | 4.24 (3.01 to 5.97) **p < .001** | |
| Repeating (≥ 3 waves) | ES-$SCZ_{75}$ = 0 | 1.0 | 1.01 (0.65 to 1.57) p = .955 | 1.33 (-0.64 to 3.29) p = .185 | 1.0 | 0.95 (0.60 to 1.51) p = 843 | 1.29 (-0.56 to 3.13) p = .171 |
| | ES-$SCZ_{75}$ = 1 | 3.42 (2.44 to 4.80) **p < .001** | 4.76 (3.00 to 7.57) **p < .001** | | 3.03 (2.14 to 4.30) **p < .001** | 4.28 (2.65 to 6.90) **p < .001** | |
| Persisting (all 4 waves) | ES-$SCZ_{75}$ = 0 | 1.0 | 0.82 (0.38 to 1.77) p = .619 | 0.37 (-2.09 to 2.83) p = .770 | 1.0 | 0.83 (0.38 to 1.82) p = .635 | -0.15 (-2.50 to 2.19) p = .898 |
| | ES-$SCZ_{75}$ = 1 | 3.48 (2.07 to 5.83) **p < .001** | 3.67 (1.82 to 7.40) **p < .001** | | 3.31 (1.90 to 5.75) **p < .001** | 2.98 (1.37 to 6.45) **p = .006** | |

Model 1: Adjusted for age and sex; Model 2: Adjusted for age, sex, family income, and parental education. P-values <.05 were bolded.

[a] Additionally adjusted for distressing PEs up to 2-year-follow-up

**Table S12.** Additive interaction between PRS$_{ice}$-SCZ$_{75}$ and ES-SCZ$_{75}$ at 2-year follow-up on distressing PEs at 3-year follow-up in the European subsample

| Distressing PEs | | Model 1 (N = 5,122) | | | Model 2 (N = 4,912) | | |
|---|---|---|---|---|---|---|---|
| | | PRS$_{ice}$-SCZ$_{75}$ = 0 OR (95% CI) | PRS$_{ice}$-SCZ$_{75}$ = 1 OR (95% CI) | RERI (95% CI) | PRS$_{ice}$-SCZ$_{75}$ = 0 OR (95% CI) | PRS$_{ice}$-SCZ$_{75}$ = 1 OR (95% CI) | RERI (95% CI) |
| [a] Past-month (3-year follow-up) | ES-SCZ$_{75}$ = 0 | 1.0 | 0.84 (0.64 to 1.12) p = .233 | 0.32 (-0.22 to 0.85) p = .250 | 1.0 | 0.84 (0.63 to 1.12) p = .233 | 0.26 (-0.27 to 0.79) p = .332 |
| | ES-SCZ$_{75}$ = 1 | 1.35 (1.08 to 1.69) **p = .008** | 1.51 (1.10 to 2.08) **p = .011** | | 1.29 (1.02 to 1.63) **p = .031** | 1.39 (1.00 to 1.94) p = .052 | |
| Lifetime (≥ 1 wave) | ES-SCZ$_{75}$ = 0 | 1.0 | 1.17 (0.95 to 1.44) p = .144 | 0.77 (-0.25 to 1.80) p = .140 | 1.0 | 1.12 (0.91 to 1.38) p = .282 | 0.73 (-0.23 to 1.68) p = .135 |
| | ES-SCZ$_{75}$ = 1 | 2.61 (2.14 to 3.18) **p < .001** | 3.55 (2.64 to 4.78) **p < .001** | | 2.39 (1.96 to 2.92) **p < .001** | 3.24 (2.40 to 4.38) **p < .001** | |
| Repeating (≥ 2 waves) | ES-SCZ$_{75}$ = 0 | 1.0 | 1.23 (0.94 to 1.62) p = .132 | 0.26 (-0.94 to 1.46) p = .672 | 1.0 | 1.16 (0.88 to 1.53) p = .298 | 0.16 (-0.96 to 1.28) p = .781 |
| | ES-SCZ$_{75}$ = 1 | 3.16 (2.48 to 4.04) **p < .001** | 3.65 (2.61 to 5.11) **p < .001** | | 2.93 (2.29 to 3.75) **p < .001** | 3.25 (2.30 to 4.58) **p < .001** | |
| Repeating (≥ 3 waves) | ES-SCZ$_{75}$ = 0 | 1.0 | 1.10 (0.71 to 1.69) p = .674 | 0.65 (-1.19 to 2.48) p = .490 | 1.0 | 1.06 (0.67 to 1.65) p = 812 | 0.81 (-0.96 to 2.59) p = .370 |
| | ES-SCZ$_{75}$ = 1 | 3.62 (2.57 to 5.09) **p < .001** | 4.36 (2.76 to 6.90) **p < .001** | | 3.19 (2.25 to 4.54) **p < .001** | 4.06 (2.52 to 6.54) **p < .001** | |
| Persisting (all 4 waves) | ES-SCZ$_{75}$ = 0 | 1.0 | 1.25 (0.62 to 2.52) p = .540 | -0.50 (-3.26 to 2.26) p = .723 | 1.0 | 1.29 (0.62 to 2.69) p = .490 | -0.71 (-3.46 to 2.04) p = .611 |
| | ES-SCZ$_{75}$ = 1 | 4.00 (2.32 to 6.88) **p < .001** | 3.74 (1.81 to 7.76) **p < .001** | | 3.75 (2.11 to 6.69) **p < .001** | 3.33 (1.51 to 7.37) **p = .003** | |

Model 1: Adjusted for age and sex; Model 2: Adjusted for age, sex, family income, and parental education. P-values <.05 were bolded.

[a] Additionally adjusted for distressing PEs up to 2-year-follow-up
